# Healthy lifestyle, genetic risk, and incidence of cancer: A prospective cohort study of 13 cancer types

**DOI:** 10.1101/2021.12.07.21267341

**Authors:** Stephanie Byrne, Terry Boyle, Muktar Ahmed, Sang Hong Lee, Beben Benyamin, Elina Hyppönen

## Abstract

**Background:** Genetic and lifestyle factors are related to the risk of cancer, but it is unclear whether a healthy lifestyle can offset genetic risk. Our aim was to investigate this for 13 cancer types using data from the UK Biobank prospective cohort.

**Methods:** In 2006-2010, participants aged 37-73 years were assessed and followed until 2015-2019. Analyses were restricted to those of European ancestries with no history of malignant cancer (n=195,822). Polygenic risk scores (PRSs) were computed for 13 cancer types and these cancers combined (‘overall cancer’), and a healthy lifestyle score was calculated from current recommendations. Relationships with cancer incidence were examined using Cox regression, adjusting for relevant confounders. Interactions between HLI and PRSs were assessed.

**Results:** There were 15,240 incident cancers during the 1,926,987 person-years of follow-up (median follow-up= 10.2 years). After adjusting for confounders, an unhealthy lifestyle was associated with a higher risk of overall cancer [lowest vs highest tertile hazard ratio (95% confidence interval) = 1.32(1.26, 1.37)] and eight cancer types. The greatest increased risks were seen for cancers of the lung [3.5(2.96,4.15)], bladder [2.03 (1.57, 2.64)], and pancreas [1.98 (1.54,2.55)]. Positive additive interactions were observed, suggesting a healthy lifestyle may partially offset genetic risk of colorectal, breast, and pancreatic cancers, and may completely offset genetic risk of lung and bladder cancers.

**Conclusions:** A healthy lifestyle is beneficial for most cancers and may offset genetic risk of some cancers. These findings have important implications for those genetically predisposed to these cancers and population strategies for cancer prevention.

## Introduction

Cancer is the second leading cause of death worldwide (1). Genetic and lifestyle factors play an important role in the aetiology of cancer. While the heritability of cancer overall has been estimated to be 33% (2), individual genetic variants typically have little impact. However, when assessed collectively using a polygenic risk score, a greater number of these genetic variants can substantially increase the likelihood of developing some cancers. Indeed, a high genetic risk (top 20%) has been reported to account for up to 30% of cases, although this varies by cancer type (3, 4).

An estimated 30-50% of all cancer cases could be prevented through healthy lifestyle behaviours, such as eating a healthy diet, being physically active, maintaining a healthy body weight, and avoiding tobacco and alcohol (1). These lifestyle factors often coexist, creating issues in estimating independent relationships with disease outcomes. Thus, research has assessed lifestyle factors collectively in a healthy lifestyle score (5). While there is strong evidence that an overall healthy lifestyle reduces risk of colorectal and breast cancers, the evidence for other cancer types is less clear (6-8).

Recent evidence suggests that genetic risk of cancer may be offset by lifestyle factors (9). The strongest evidence has been obtained from studies on breast and colorectal cancer, which suggest a healthy lifestyle may reduce the risk regardless of genetic risk (10-12), and may even be of greater benefit for those at high genetic risk (13, 14). However, these relationships have not been explored for other cancer types. In this study, we investigate whether a healthy lifestyle can offset genetic risk and assess associations with 13 different types of cancer - bladder, breast, colorectal, kidney, lymphocytic leukaemia, lung, melanoma, non-Hodgkin lymphoma (NHL), oral cavity/pharyngeal, ovarian, pancreatic, prostate, and uterine cancer.

## Methods

### Study design and study population

This prospective cohort study utilizes data from the UK Biobank, a large population-based cohort of over 500,000 adults aged 37-73 years and living in the United Kingdom at the time of the initial assessment in 2006-2010 (15). Participants attended one of 22 assessment centres across England, Scotland and Wales and completed an assessment including a questionnaire, anthropometric measures, and collection of biological samples. UK Biobank was approved by the National Health Service North West Multi-centre Research Ethics Committee (11/NW/0382), the National Information Governance Board for Health and Social Care in England and Wales, and the Community Health Index Advisory Group in Scotland. All participants provided written informed consent.

To reduce the effects of population stratification, our analyses were restricted to participants of white European ancestries, and we included participants with no history of any malignant cancer at baseline and sufficient data available for the lifestyle factors and other covariates of interest (Figure 1).

**Figure 1:**
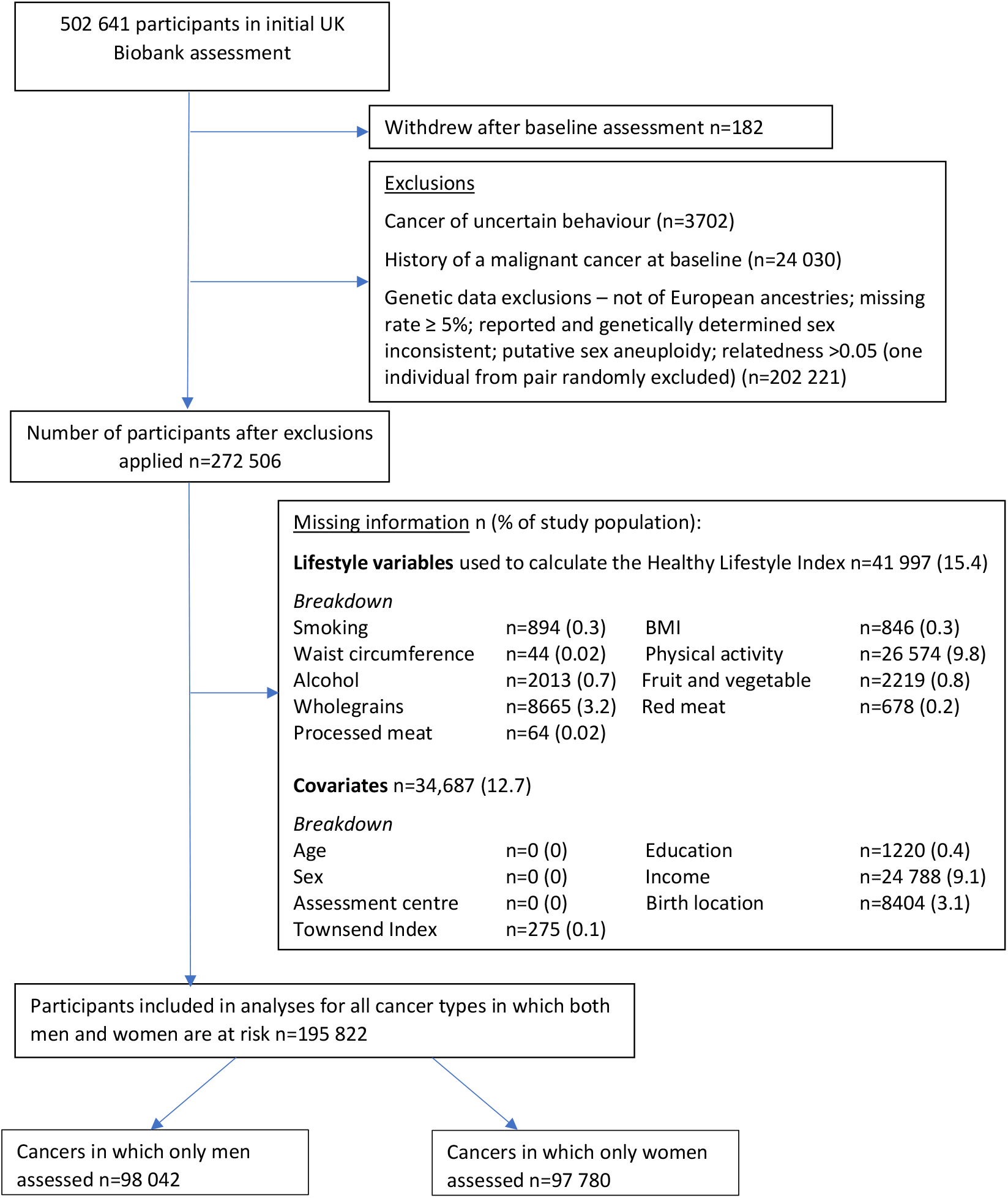
Ascertainment of the study sample included in the analyses

### Polygenic risk scores (PRSs)

We calculated PRSs for each participant for 13 cancer types based on single-nucleotide polymorphisms (SNPs) selected by Graff et al (3). Briefly, they identified a set of independent SNPs associated with cancer risk for each cancer type from previously published cancer genome-wide association studies. SNP selection was restricted to those that are common and available in the UK Biobank data. SNPs with the strongest associations with the broadest phenotype were preferentially selected, with an overall linkage disequilibrium r^2^ <0.3 to ensure independence (see Supplementary Table S1 for the number of SNPs used in the PRS calculations. See Graff et al for a full list of genetic variants (3)).

We calculated PRSs as the sum of the number of risk alleles multiplied by the log of the odds ratio (OR) for each SNP, as implemented in PLINK software (16) using the following formula:

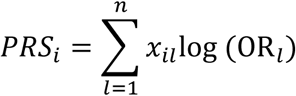

where *n* = the number of SNPs in the set of independent SNPs for that cancer type, *x*_*il*_ is the number of risk alleles (0, 1 or 2) for the *i*th individual at the *l*th SNP, and *OR*_*l*_ is the estimated OR for the *l*th SNP using a logistic regression.

Each PRS was z-standardized, with higher scores representing a greater genetic risk, and then categorized as low (lowest tertile), intermediate (middle tertile), and high (highest tertile) risk.

A PRS for overall cancer was also derived as the sum of the z-standardized PRSs for the 13 cancer types, each weighted according to the distribution of incident cases of these cancers reported by Cancer Research UK in 2017 (https://www.cancerresearchuk.org/health-professional/cancer-statistics/incidence#heading-Four).

### Healthy lifestyle index

A healthy lifestyle index (HLI) was constructed based on the World Cancer Research Fund/American Institute of Cancer Research (WCRF/AICR) cancer prevention recommendations and standardized scoring system (full details of the calculation of the HLI are in Supplementary Table S2) (5). Baseline anthropometric and touchscreen questionnaire information was available for five of the eight recommendations included in the 2018 WCRF/AICR score (healthy weight; physical activity; wholegrain, fruit and vegetable intake; red and processed meat intake; alcohol consumption). Smoking status (categorized as never, former, or current smoking) was also included in the HLI, given its relevance as a modifiable lifestyle risk factor for multiple cancers. The HLI ranged from 0 to 6, with higher scores indicating a greater adherence to a healthy lifestyle. The HLI was then split into tertiles and categorized as unfavourable (HLI range 0-3), intermediate (HLI range 3.25-3.75), and favourable (HLI range 4-6) lifestyle.

### Cancer outcome ascertainment

Malignant cancer diagnoses for each cancer type were identified using the ninth and tenth revisions of the International Classification of Diseases (ICD-9 and ICD-10) codes, phenotype, and tumour behaviour information obtained through linkage to national cancer registries in England, Wales, and Scotland. Incident cancer events were those diagnosed after baseline assessment. Cancer information was complete up to 31 July 2019 for England and Wales, and 31 October 2015 for Scotland. Hence, follow-up time was defined as being from the date of baseline assessment to the earliest of date of diagnosis, date of death (obtained via linkage to death registries) or 31 July 2019 for England and Wales residents and 31 October 2015 for Scotland residents.

### Statistical analysis

Baseline characteristics were summarized for the total study population, those diagnosed with any of the 13 cancers examined (referred to as “overall cancer”), and individually for each cancer type. Characteristics were summarized as percentages for all variables for greater interpretability.

Cox proportional hazard regression models were used to investigate associations between PRS categories, HLI categories, and incidence of overall cancer and separately for each cancer. Incidence of cancers that are sex-specific were assessed in the relevant sex only (female cancers – ovarian, uterine, breast; male cancers – prostate). Breast cancer analyses were further restricted to post-menopausal women only (self-reported at baseline). All models were adjusted for age at baseline (continuous); sex (where relevant); assessment centre; socioeconomic status (Townsend Index - continuous); education (none listed, GCSE/CSE or equivalent, A levels or equivalent, college/university or other professional training); household income (<18,000, 18,000 to 30,999, 31,000 to 51,999, 52,000 to 100,000, >100,000); birth location (north south coordinate and east west coordinate – both in deciles), and population stratification measured by the first 40 principal components. Trend p-values were calculated by using the PRS and HLI categorical variables as pseudo-continuous variables in the models. Subsequent analyses adjusting for additional covariates specific to each cancer type were also performed (see Supplementary Table S3 for a list of the additional covariates and definitions). Additional analyses exploring the risk of cancer in the top 5% of PRS were conducted using Cox regression with the bottom tertile of PRS as the reference and adjusting for the same covariates.

For each cancer outcome, we investigated multiplicative interactions between lifestyle and genetic risk by using a likelihood ratio test to compare Cox models with and without an interaction term between PRS categories and HLI categories. Additive interactions were assessed by the relative excess risk due to interaction (RERI), which were calculated from the Cox models with the interaction term (17). Observed interactions were further explored visually using a forest plot produced from a multivariable Cox proportional hazards model with a combined genetic and lifestyle risk variable (9 categories with low genetic risk and favourable lifestyle as the reference). As smoking is a strong risk factor for lung cancer, we performed sensitivity analyses removing smoking from the HLI and including it as a confounder for this outcome. Proportionality of hazards assumptions were verified using Schoenfeld residuals and visual inspection of log-log and residuals plots. Two-sided p-values less than or equal to 0.05 were considered as evidence of an association. A p-value threshold of 0.0036 (calculated as 0.05/number of cancer outcomes) was applied to interaction models to account for multiple testing. All analyses were performed using Stata SE version 16 (StataCorp, College Station, TX). Results are presented in order of power determined from the number of cases for each cancer type.

## Results

A summary of the baseline characteristics of the 195 822 participants included in the analysis, and for those diagnosed with any of the 13 types of cancer during the follow up period, are provided in Table 1. Over the 1 926 987 person-years of follow up (median [interquartile range] length of follow up = 10.2 [9.4-10.9] years), there was a total of 15 240 incident cases of the 13 cancer types of interest. The cancers with the highest number of incident cases were prostate cancer, colorectal cancer, and post-menopausal breast cancer. Generally, risk of cancer was higher in men, those of older age, and those who have lower levels of education and income. An unfavourable lifestyle and a high genetic predisposition were also associated with a higher incidence of most, but not all, cancers.

**Table 1.** Baseline characteristics of the total study population and of incident cases of each cancer type

| Characteristic | Total Study population | Overall cancer | Prostate cancer | Colorectal cancer | Post-menopausal breast cancer | Lung cancer | Melanoma | Non-Hodgkin lymphoma | Kidney cancer | Uterine cancer | Pancreatic cancer | Bladder cancer | Oral cavity/ pharyngeal cancer | Ovarian cancer | Lymphocytic leukaemia |
| --- | --- | --- | --- | --- | --- | --- | --- | --- | --- | --- | --- | --- | --- | --- | --- |
| N | 195 822 | 15 240 | 4476 | 2150 | 1990 | 1256 | 1200 | 721 | 547 | 494 | 451 | 424 | 415 | 315 | 246 |
| Age at baseline assessment (%)<sup>a</sup> |  |  |  |  |  |  |  |  |  |  |  |  |  |  |  |
| 50 years and under | 27.0 | 11.5 | 4.8 | 9.8 | 2.1 | 5.3 | 17.5 | 9.2 | 8.4 | 11.1 | 6.7 | 3.3 | 13.7 | 20.0 | 5.7 |
| 51 - 60 years | 36.7 | 33.1 | 29.4 | 32.5 | 45.5 | 30.3 | 33.5 | 32.3 | 33.8 | 41.9 | 29.7 | 23.8 | 40.5 | 30.8 | 31.7 |
| 61 years and over | 36.4 | 55.4 | 65.8 | 57.7 | 52.4 | 64.4 | 49.0 | 58.5 | 57.8 | 47.0 | 63.6 | 72.9 | 45.8 | 49.2 | 62.6 |
| Female (%) | 49.9 | 43.5 | 0 | 38.8 | 100 | 44.2 | 45.7 | 41.3 | 33.8 | 100 | 41.0 | 17.5* | 29.2 | 100 | 29.7 |
| Townsend Index (%)<sup>a</sup> |  |  |  |  |  |  |  |  |  |  |  |  |  |  |  |
| quintile 1 | 22.5 | 22.6 | 24.6 | 22.8 | 22.5 | 15.2 | 26.0 | 25.4 | 24.3 | 17.0 | 22.4 | 21.0 | 16.4 | 20.6 | 24.0 |
| quintile 2 | 21.6 | 22.3 | 23.2 | 23.7 | 21.8 | 19.2 | 25.0 | 19.7 | 21.0 | 23.9 | 23.1 | 24.5 | 22.7 | 24.8 | 17.5 |
| quintile 3 | 20.7 | 20.6 | 21.6 | 19.7 | 20.5 | 17.8 | 19.9 | 21.8 | 24.5 | 19.6 | 22.6 | 19.6 | 17.4 | 19.1 | 26.8 |
| quintile 4 | 19.4 | 18.9 | 18.1 | 18.0 | 20.9 | 20.1 | 17.5 | 18.9 | 15.7 | 21.1 | 15.5 | 18.2 | 21.5 | 20.6 | 17.5 |
| quintile 5 | 15.8 | 15.6 | 12.6 | 15.8 | 14.4 | 27.7 | 11.6 | 14.3 | 14.4 | 18.4 | 16.4 | 16.8 | 22.2 | 14.9 | 14.2 |
| Education (%)<sup>a</sup> |  |  |  |  |  |  |  |  |  |  |  |  |  |  |  |
| None listed | 12.6 | 16.3 | 15.5 | 15.7 | 15.1 | 30.0 | 13.2 | 18.5 | 18.1 | 13.4 | 19.3 | 21.0 | 19.0 | 15.6 | 20.7 |
| GCSE/CSE or equivalent | 27.1 | 25.5 | 21.6 | 26.3 | 29.2 | 25.1 | 27.7 | 22.2 | 28.0 | 31.4 | 22.4 | 27.1 | 26.3 | 31.4 | 24.8 |
| A levels or equivalent | 12.3 | 11.3 | 10.5 | 11.8 | 12.2 | 10.0 | 10.9 | 10.8 | 11.9 | 10.5 | 9.5 | 8.5 | 12.1 | 11.4 | 8.9 |
| College/University or other professional training | 48.0 | 47.0 | 52.4 | 46.2 | 43.6 | 35.0 | 48.3 | 48.5 | 42.1 | 44.7 | 48.8 | 43.4 | 42.7 | 41.6 | 45.5 |
| Average total household income before tax (%)<sup>a</sup> |  |  |  |  |  |  |  |  |  |  |  |  |  |  |  |
| Less than 18,000 £ | 19.4 | 23.8 | 20.4 | 23.8 | 25.7 | 41.2 | 18.3 | 22.6 | 24.7 | 25.3 | 27.7 | 29.0 | 28.9 | 27.9 | 30.1 |
| 18,000 to 30,999 £ | 25.2 | 28.4 | 27.7 | 29.7 | 31.7 | 30.1 | 23.8 | 32.6 | 29.8 | 32.0 | 27.3 | 30.7 | 25.8 | 31.1 | 26.4 |
| 31,000 to 51,999 £ | 27.2 | 25.6 | 26.8 | 25.6 | 24.3 | 18.4 | 27.8 | 26.9 | 25.2 | 24.3 | 25.3 | 22.6 | 25.1 | 24.4 | 26.0 |
| 52,000 to 100,000 £ | 22.4 | 17.4 | 19.6 | 16.0 | 15.2 | 8.1 | 24.3 | 13.5 | 16.3 | 15.2 | 14.6 | 13.9 | 16.6 | 12.4 | 15.0 |
| Greater than 100,000 £ | 5.9 | 4.8 | 5.5 | 4.9 | 3.1 | 2.2 | 5.8 | 4.4 | 4.0 | 3.2 | 5.1 | 3.8 | 3.6 | 4.1 | 2.4 |
| HLI (%)<sup>a</sup> |  |  |  |  |  |  |  |  |  |  |  |  |  |  |  |
| Unfavourable | 36.6 | 42.0 | 41.0 | 44.9 | 33.6 | 63.1 | 34.8 | 38.0 | 46.6 | 38.3 | 48.3 | 55.9 | 50.6 | 30.5 | 38.2 |
| Intermediate | 32.4 | 32.0 | 32.2 | 30.6 | 36.9 | 23.5 | 35.5 | 31.9 | 35.1 | 34.0 | 32.2 | 25.9 | 28.9 | 35.9 | 33.3 |
| Favourable | 31.0 | 26.0 | 26.8 | 24.5 | 29.6 | 13.4 | 29.8 | 30.1 | 18.3 | 27.7 | 19.5 | 18.2 | 20.5 | 33.7 | 28.5 |
| PRS (%)<sup>a</sup> |  |  |  |  |  |  |  |  |  |  |  |  |  |  |  |
| Low genetic risk | - | 30.5 | 24.3 | 22.4 | 24.9 | 25.5 | 23.0 | 27.7 | 27.8 | 22.3 | 20.6 | 24.3 | 32.8 | 26.7 | 22.0 |
| Intermediate genetic risk | - | 33.3 | 31.2 | 32.9 | 33.1 | 32.9 | 31.2 | 33.3 | 32.2 | 36.2 | 33.3 | 36.6 | 33.0 | 34.0 | 24.8 |
| High genetic risk | - | 36.3 | 44.5 | 44.7 | 42.0 | 41.6 | 45.8 | 39.0 | 40.0 | 41.5 | 46.1 | 39.2 | 34.2 | 39.4 | 53.3 |

Abbreviations: GCSE – General Certificate of Secondary Education, CSE – Certificate of Secondary education, HLI – Healthy Lifestyle Index, PRS –
Polygenic Risk Score
<sup>a</sup> Percentages may not add to 100% due to rounding

After adjusting for confounders, an unhealthy lifestyle was associated with a higher risk of overall cancer [lowest vs highest tertile of HLI hazard ratio (HR) 95% confidence interval (95%CI) = 1.32 (1.26, 1.37)], colorectal cancer [1.42 (1.28, 1.59)], post-menopausal breast cancer [1.42 (1.27, 1.59)], lung cancer [3.50 (2.96, 4.15)], kidney cancer [1.91 (1.51, 2.42)], uterine cancer [1.63 (1.31, 2.04)], pancreatic [1.98 (1.54, 2.55)], bladder cancer [2.03 (1.57, 2.64)], and oral cavity/pharyngeal cancer [1.69 (1.31, 2.18)] (Figure 2). There was no association between HLI and melanoma, NHL, ovarian cancer, and lymphocytic leukaemia. A weak association in the opposite direction was observed between HLI and prostate cancer risk [0.86 (0.79, 0.94)] (Figure 2). Further adjustment for other potential confounders specific to cancer type had little effect on the estimates presented (Supplementary Table S4).

**Figure 2:**
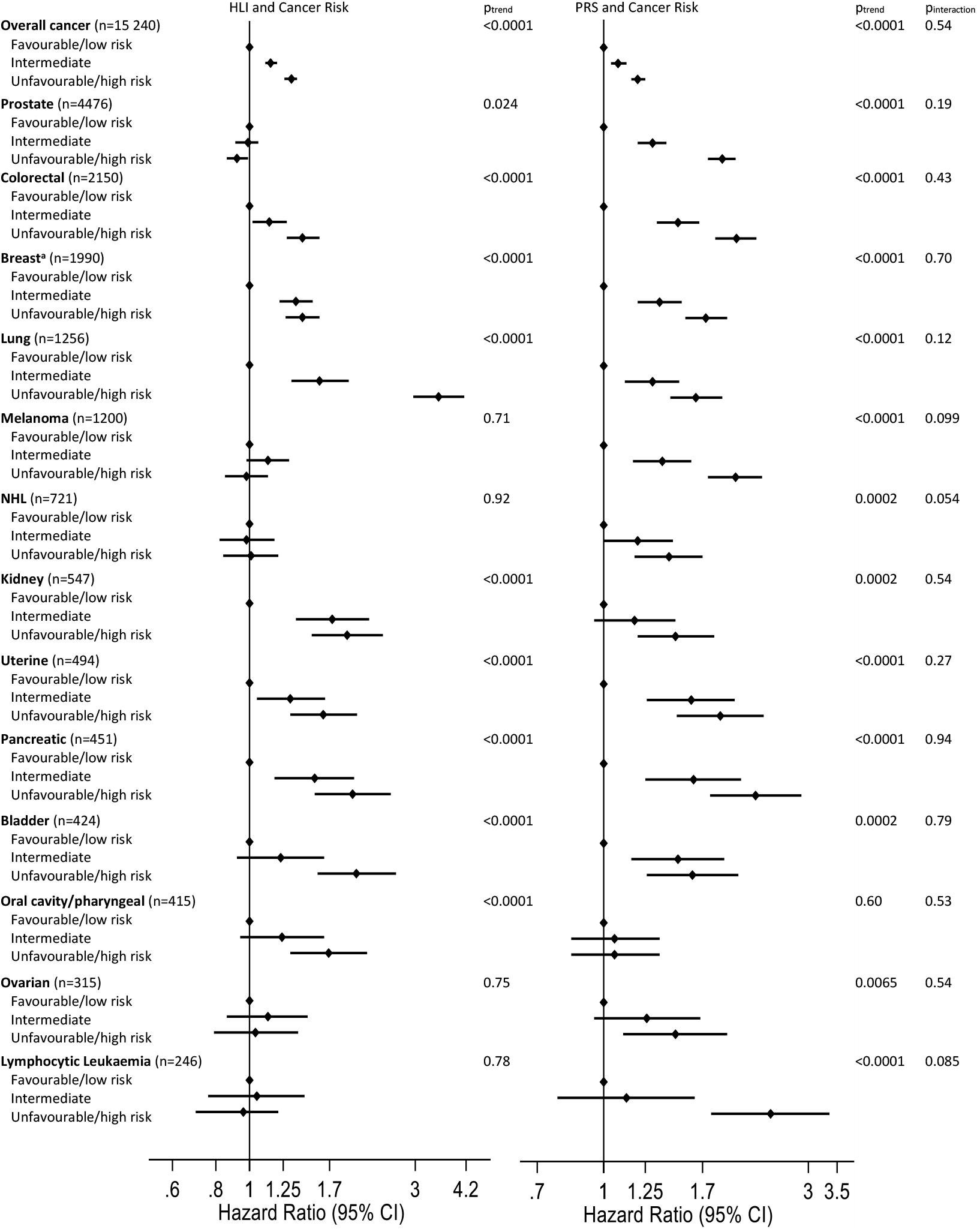
Adjusted associations between the Healthy Lifestyle Index, Polygenic Risk Score, and cancer Cox proportional hazards regression adjusted for age at baseline, sex (where relevant), assessment centre, 40 principal components of ancestries, Townsend Index, education, birth location, and income; overall cancer = overall incident cases of the 13 cancer types assessed in this study; n=number of cases; ^a^Post-menopausal breast cancer

A higher PRS was associated with higher risk of overall cancer and all cancer types assessed, except oral cavity/pharyngeal cancer (Figure 2). The cancers with the greatest increased risk in those with a high PRS compared to those with a low PRS were colorectal cancer [2.04 (1.82, 2.27)], melanoma [2.03 (1.75, 2.34), pancreatic cancer [2.26 (1.77, 2.89)], and lymphocytic leukaemia [2.45 (1.78, 3.36)]. Again, further adjustment for other cancer-specific potential confounders had little impact on the hazard ratios (Supplementary Table S4). Further exploration of genetic risk found those in the top 5% of PRS for prostate, colorectal, melanoma, and pancreatic cancers had an estimated 2.5-fold, 2.5-fold, 3-fold, and 4-fold increased risk, respectively (Table 2).

**Table 2.** Risk of cancer for those in the top 5% of polygenic risk score

| Cancer Type | Top 5% of polygenic risk score |  |  |
| --- | --- | --- | --- |
|  | Number of cases | HR 95%CI<sup>a</sup> | p-value |
| Overall cancer | 896 | 1.33 (1.24, 1.43) | <0.0001 |
| Prostate cancer | 383 | 2.53 (2.25, 2.84) | <0.0001 |
| Colorectal cancer | 180 | 2.52 (2.13, 3.00) | <0.0001 |
| Post-menopausal breast cancer | 225 | 1.92 (1.59, 2.32) | <0.0001 |
| Lung cancer | 104 | 2.22 (1.78, 2.77) | <0.0001 |
| Melanoma | 121 | 3.01 (2.43, 3.74) | <0.0001 |
| Non-Hodgkin's lymphoma | 61 | 2.06 (1.54, 2.74) | <0.0001 |
| Kidney cancer | 28 | 1.25 (0.83, 1.87) | 0.28 |
| Uterine cancer | 35 | 2.22 (1.52, 3.25) | <0.0001 |
| Pancreatic cancer | 57 | 4.14 (2.97, 5.76) | <0.0001 |
| Bladder cancer | 31 | 1.99 (1.33, 2.98) | 0.0008 |
| Oral cavity/pharyngeal cancer | 26 | 1.31 (0.86, 1.99) | 0.21 |
| Ovarian cancer | 20 | 1.55 (0.95, 2.53) | 0.078 |
| Lymphocytic leukaemia | 24 | 3.00 (1.85, 4.85) | <0.0001 |

<sup>a</sup> reference category = bottom tertile of polygenic risk score
All models adjusted for healthy lifestyle index category, age at baseline, sex, assessment centre, 40 principal components of ancestries,

There were no multiplicative interactions observed between HLI and PRS for any of the cancer outcomes (p>0.05 for all outcomes, Figure 2). However, additive interactions were observed for colorectal, breast, lung, pancreatic, and bladder cancers (Supplementary Table S5). Forest plots of risk with a combined lifestyle/genetic risk variable were constructed to visually depict these interactions. These plots show a greater increased risk with a less favourable lifestyle in those with a higher genetic risk for these cancers, with a favourable lifestyle completely offsetting the increased risk from genetic factors for lung and bladder cancers (Figures 3).

**Figure 3:**
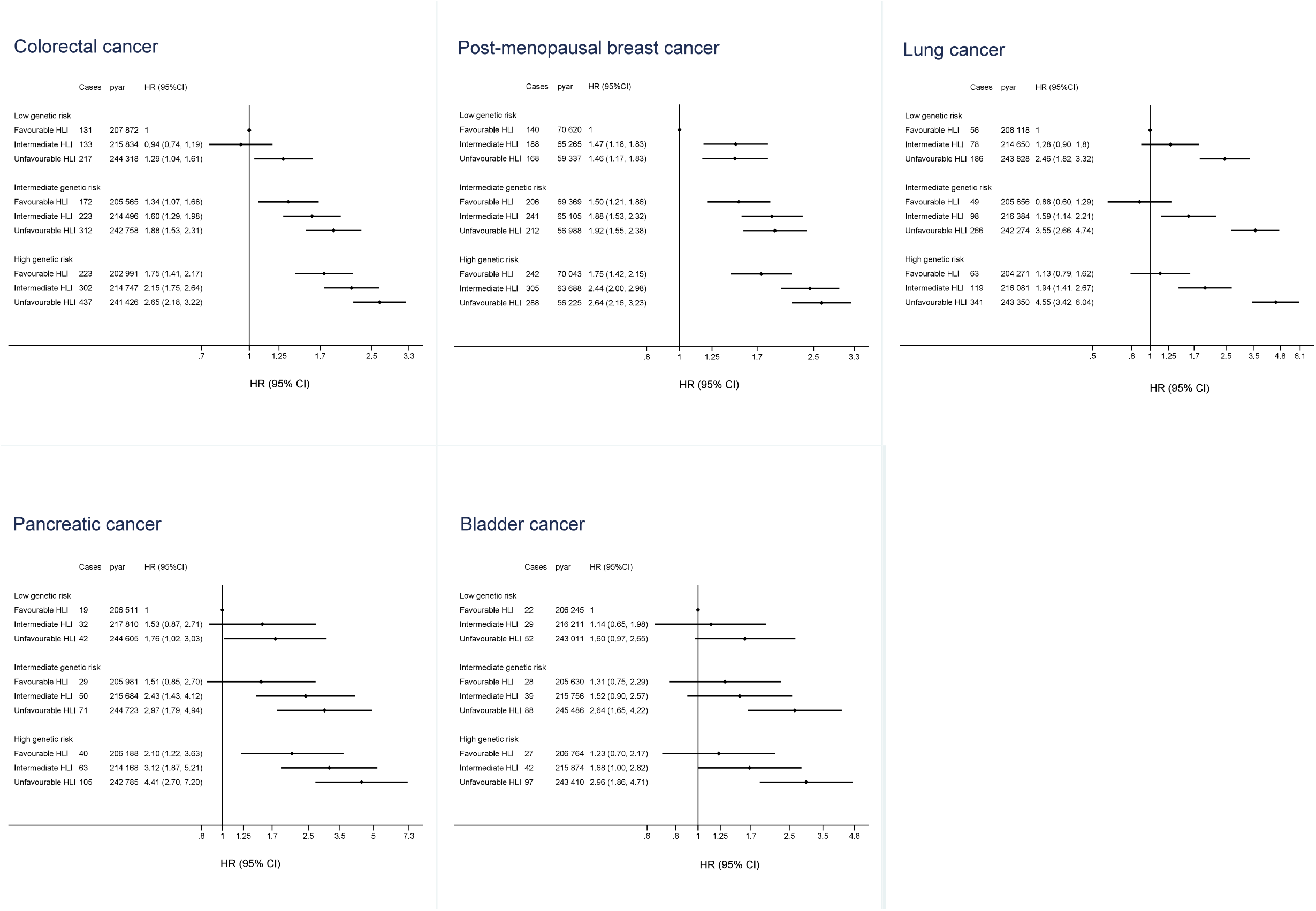
Risk of colorectal, post-menopausal breast, lung, pancreatic and bladder cancers
according to genetic risk and lifestyle Cox proportional hazards regression adjusted for age at baseline, sex (where relevant), assessment centre, 40 principal components of ancestries, Townsend Index, education, birth location, and income; HLI=Healthy Lifestyle Index; pyar=personyears at risk; HR (95%CI) = Hazard ratio (95% confidence interval)

We further investigated the association between lifestyle and lung cancer by removing smoking status from the HLI and including it as a confounder. In this analysis, an unhealthy lifestyle was associated with a higher risk of lung cancer [lowest vs highest tertile of HLI without smoking HR (95%CI) = 1.23 (1.06, 1.42), p=0.0055]. There was no multiplicative interaction between HLI (without smoking) and smoking status for lung cancer (p=0.393). The additive interaction between HLI and PRS was attenuated [unfavourable lifestyle/high genetic risk vs favourable lifestyle/low genetic risk RERI=0.44 (0.04, 0.85) p=0.033; p_trend_=0.021].

## Discussion

In this study, an unfavourable lifestyle and a high genetic risk were independently associated with an increased risk of several types of cancer. There were no multiplicative interactions between lifestyle and genetic risk, but additive interactions were observed. These findings suggest a healthy lifestyle may be of greater benefit in those with a high genetic susceptibility to colorectal, breast, and pancreatic cancers, and may completely offset genetic risk for lung and bladder cancers. Our findings are consistent with the limited comparable research for breast (10, 13) and colorectal cancers (11, 12, 14) and is novel for the other cancer types examined here. Communicating these results to groups with a high genetic risk of cancer may help alleviate any distress experienced due to awareness of their increased risk, and the appreciation of some control over the genetic risk is likely to be empowering and potentially supportive of positive behavioural changes.

A higher genetic risk of cancer was associated with an increased risk of overall cancer and 12 cancer types – prostate, colorectal, breast (post-menopause), lung, melanoma, NHL, kidney, uterine, pancreatic, bladder, ovarian, and lymphocytic leukaemia. The highest genetic risk was observed for pancreatic cancer, with those in the top 5% having a 4-fold higher risk compared to those in the bottom tertile of genetic risk. This increase is similar in magnitude to *BRCA1* and *BRCA2* gene variants and breast cancer risk, which are far less common (prevalence of 0.2-0.3%) and trigger more frequent cancer screening (18). Pancreatic cancer is one of the leading causes of cancer death that is typically diagnosed at a late stage when the 5-year survival rate is less than 10% (19). Screening for pancreatic cancer is recommended for those deemed high-risk (20), and our results suggest a PRS could be used as an additional tool to assist in the identification of those at high risk. We also found that those with a poor lifestyle had an increased risk of pancreatic cancer and this increase in risk was greater in those with a high genetic risk. Therefore, those with a high genetic susceptibility to pancreatic cancer may benefit more from a healthy lifestyle than their low genetic risk counterparts. This adds value to the identification of high-risk groups and highlights opportunities for prevention intervention programs.

An unhealthy lifestyle was associated with a higher risk of overall cancer and eight cancer types - colorectal, postmenopausal breast, lung, kidney, uterine, pancreatic, bladder, and oral cavity/pharyngeal cancers. These findings are consistent with previously published research for breast and colorectal cancers and add to the limited and inconclusive research on overall healthy lifestyle and other cancers (6, 7). In addition to pancreatic cancer, we also observed additive interactions for colorectal, breast, lung, and bladder cancers, suggesting the harmful effects of a poor lifestyle are higher with increased genetic risk and a healthy lifestyle may partially or fully offset genetic risk of these cancers. These findings reinforce the current public health message that living a healthy lifestyle including not smoking, avoiding alcohol, consuming a healthy diet, maintaining a healthy body weight, and engaging in regular physical activity, reduces cancer risk.

We found no relationship between a healthy lifestyle and risk of melanoma, NHL, ovarian cancer, and lymphocytic leukaemia. These results were not unexpected; none of the lifestyle factors included in the HLI have been conclusively linked with the risk of lymphocytic leukaemia, melanoma, or NHL, and only obesity is positively associated with ovarian cancer risk (8). For melanoma, although evidence indicates higher alcohol intake may increase risk, greater physical activity levels have also been associated with increased risk, which is likely related to higher sun exposure levels (21, 22).

We found a healthy lifestyle was associated with a higher risk of prostate cancer. This finding is inconsistent with the majority of previous research suggesting either a negative association or no relationship for combined or individual components of a healthy lifestyle (6, 23-28), and may be the result of healthy volunteer bias.

This large prospective cohort study has some limitations. The length of the follow up and age range of the study population has limited the number of incident cancer cases. Consequently, some statistical models may not have been adequately powered to observe a modifying effect of genetic risk on healthy lifestyle-cancer relationships. There is likely to be measurement errors in the components of the lifestyle score as they are almost all measured via self-report; however, this is likely to attenuate the findings to the null. We also cannot rule out the possibility of residual confounding, although we adjusted for multiple covariates. The measure of genetic risk was limited by the SNPs included in the PRS, which may not be exhaustive. Lastly, this study was conducted in adults of European ancestries, so the relevance of these findings to populations of other ethnicities is unclear.

In summary, our findings indicate individuals can reduce their cancer risk by adhering to the WCRF/AICR healthy lifestyle recommendations and, for some cancers, those who are genetically susceptible to cancer can partially or fully offset their increased risk by living healthily. Our findings also highlight the potential benefit of using a PRS to identify those with a high genetic risk of pancreatic cancer, and possibly other cancers, as PRSs are further refined. PRSs could be used in conjunction with other tools to increase the accuracy of identifying high-risk individuals who may then undergo regular screening and be targeted by prevention intervention strategies. This prospect requires further investigation using cohort studies conducted across various ethnic populations.

## Data Availability

This research utilizes data from the UK Biobank resource (application number 20175) which is available directly from the UK Biobank upon submission of a data request proposal. See https://www.ukbiobank.ac.uk.

## Declarations

### Ethics approval

This study is a secondary analysis of UK Biobank data. The UK Biobank cohort study was approved by the National Health Service North West Multi-centre Research Ethics Committee (11/NW/0382), the National Information Governance Board for Health and Social Care in England and Wales, and the Community Health Index Advisory Group in Scotland. All participants provided written informed consent.

### Author contributions

Data acquisition: Lee, Hypponen; Concept and design: Byrne, Boyle, Benyamin, Hypponen; Data analysis: Byrne, Boyle, Lee, Benyamin; Data management: Byrne, Ahmed, Benyamin; Drafting of the manuscript: Byrne; Obtaining funding: Boyle, Benyamin, Hypponen. All authors were involved in the interpretation of the results and the review and final approval of the manuscript. Drs Byrne and Boyle had full access to all the data and take full responsibility for the integrity of the data and the accuracy of the analysis.

### Supplementary material

Supplementary material is available online.

## References

1. World Health Organisation. Cancer. 2021. https://www.who.int/health-topics/cancer (18 November 2021, date last accessed).

2. Mucci LA, Hjelmborg JB, Harris JR, et al. Familial Risk and Heritability of Cancer Among Twins in Nordic Countries. JAMA 2016;315(1):68–76.

3. Graff R, Cavazos T, Thai K, et al. Cross-cancer evaluation of polygenic risk scores for 16 cancer types in two large cohorts. Nat Commun 2021;12(1):970.

4. Kachuri L, Graff RE, Smith-Byrne K, et al. Pan-cancer analysis demonstrates that integrating polygenic risk scores with modifiable risk factors improves risk prediction. Nat Commun 2020;11(1):6084.

5. Shams-White MM, Brockton NT, Mitrou P, et al. Operationalizing the 2018 World Cancer Research Fund/American Institute for Cancer Research (WCRF/AICR) Cancer Prevention Recommendations: A Standardized Scoring System. Nutrients 2019;11(7):1572.

6. Solans M, Chan DSM, Mitrou P, Norat T, Romaguera D. A systematic review and meta-analysis of the 2007 WCRF/AICR score in relation to cancer-related health outcomes. Ann Oncol 2020;31(3):352–68.

7. Zhang YB, Pan XF, Chen J, et al. Combined lifestyle factors, incident cancer, and cancer mortality: a systematic review and meta-analysis of prospective cohort studies. Br J Cancer 2020;122(7):1085–93.

8. World Cancer Research Fund/American Institute for Cancer Research. Diet, Nutrition, Physical Activity and Cancer: a Global Perspective. 2018. https://www.wcrf.org/diet-and-cancer/ (4 March 2021, date last accessed).

9. Zhu M, Wang T, Huang Y, et al. Genetic Risk for Overall Cancer and the Benefit of Adherence to a Healthy Lifestyle. Cancer Res 2021;81(17):4618–27.

10. Al Ajmi K, Lophatananon A, Mekli K, Ollier W, Muir KR. Association of Nongenetic Factors With Breast Cancer Risk in Genetically Predisposed Groups of Women in the UK Biobank Cohort. JAMA Netw Open 2020;3(4):e203760.

11. Cho YA, Lee J, Oh JH, et al. Genetic Risk Score, Combined Lifestyle Factors and Risk of Colorectal Cancer. Cancer Res Treat 2019;51(3):1033–40.

12. Carr PR, Weigl K, Jansen L, et al. Healthy Lifestyle Factors Associated With Lower Risk of Colorectal Cancer Irrespective of Genetic Risk. Gastroenterology 2018;155(6):1805-15.e5.

13. Arthur RS, Wang T, Xue X, Kamensky V, Rohan TE. Genetic Factors, Adherence to Healthy Lifestyle Behavior, and Risk of Invasive Breast Cancer Among Women in the UK Biobank. J Natl Cancer Inst 2020;112(9):893–901.

14. Choi J, Jia G, Wen W, Shu XO, Zheng W. Healthy lifestyles, genetic modifiers, and colorectal cancer risk: a prospective cohort study in the UK Biobank. Am J Clin Nutr 2021;113(4):810–820.

15. Sudlow C, Gallacher J, Allen N, et al. UK biobank: an open access resource for identifying the causes of a wide range of complex diseases of middle and old age. PLoS Med 2015;12(3):e1001779.

16. Purcell S, Neale B, Todd-Brown K, et al. PLINK: a tool set for whole-genome association and population-based linkage analyses. Am J Hum Genet 2007;81(3):559–75.

17. VanderWeele TJ, Knol MJ. A Tutorial on Interaction. Epidemiol Methods 2014;3(1):33–72.

18. National Cancer Institute. BRCA Gene Mutations: Cancer Risk and Genetic Testing. 2020. https://www.cancer.gov/about-cancer/causes-prevention/genetics/brca-fact-sheet (3 June 2021, date last accessed).

19. Klein AP. Pancreatic cancer epidemiology: understanding the role of lifestyle and inherited risk factors. Nat Rev Gastroenterol Hepatol 2021;18(1):493–502.

20. Goggins M, Overbeek KA, Brand R, et al. Management of patients with increased risk for familial pancreatic cancer: updated recommendations from the International Cancer of the Pancreas Screening (CAPS) Consortium. Gut 2020;69(1):7–17.

21. Moore SC, Lee I-M, Weiderpass E, et al. Association of Leisure-Time Physical Activity With Risk of 26 Types of Cancer in 1.44 Million Adults. JAMA Intern Med 2016;176(6):816–25.

22. Holman DM, Berkowitz Z, Guy GP, Hartman AM, Perna FM. The association between demographic and behavioral characteristics and sunburn among U.S. adults — National Health Interview Survey, 2010. Prev Med 2014;63:6–12.

23. Olmedo-Requena R, Lozano-Lorca M, Salcedo-Bellido I, et al. Compliance with the 2018 World Cancer Research Fund/American Institute for Cancer Research Cancer Prevention Recommendations and Prostate Cancer. Nutrients 2020;12(3):768.

24. Romaguera D, Gracia-Lavedan E, Molinuevo A, et al. Adherence to nutrition-based cancer prevention guidelines and breast, prostate and colorectal cancer risk in the MCC-Spain case-control study. Int J Cancer 2017;141(1):83–93.

25. Lavalette C, Adjibade M, Srour B, et al. Cancer-Specific and General Nutritional Scores and Cancer Risk: Results from the Prospective NutriNet-Sante Cohort. Cancer Res 2018;78(15):4427–35.

26. Pernar CH, Ebot EM, Wilson KM, Mucci LA. The Epidemiology of Prostate Cancer. Cold Spring Harb Perspect Med 2018;8(12) :a030361.

27. Schulpen M, van den Brandt PA. Adherence to the Mediterranean Diet and Risks of Prostate and Bladder Cancer in the Netherlands Cohort Study. Cancer Epidemiol Biomarkers Prev 2019;28(9):1480–8.

28. Solans M, Romaguera D, Gracia-Lavedan E, et al. Adherence to the 2018 WCRF/AICR cancer prevention guidelines and chronic lymphocytic leukemia in the MCC-Spain study. Cancer Epidemiol 2020;64(1):101629.

