## Supplementary material for "Healthy lifestyle, genetic risk, and incidence of cancer: A prospective cohort study of 13 cancer types"

### Online Supplement

#### Supplementary Tables

Table S1 – Number of SNPs used to calculate the PRS for each cancer type

Table S2 - Definition of lifestyle factors

Table S3 – Definition of additional covariates included in further statistical models

Table S4 - Adjusted associations between the Healthy Lifestyle Index, Polygenic Risk Score, and cancer risk

Table S5 – Relative excess risk due to interaction (RERI) for additive interactions between Healthy Lifestyle Index and Polygenic Risk Score for overall cancer and 13 cancer types

**Table S1 – Number of SNPs used to calculate the PRS for each cancer type**

| Cancer Type | Number of SNPs used in PRS calculation |
|---|---|
| Prostate | 161 |
| Colorectal | 103 |
| Breast | 187 |
| Lung | 109 |
| Melanoma | 24 |
| Non-Hodgkin lymphoma | 19 |
| Kidney | 19 |
| Uterine | 9 |
| Pancreatic | 22 |
| Bladder | 15 |
| Oral cavity/pharynx | 14 |
| Ovarian | 36 |
| Lymphocytic leukaemia | 75 |

SNP – single nucleotide polymorphism; PRS – polygenic risk score

#### Table S2 - Definition of lifestyle factors

| Lifestyle factor | Measure | Classification | |
|---|---|---|---|
| Be a healthy weight | BMI (kg/m2) | 18.5 to <25 | 0.5 |
| | | 25 to <30 | 0.25 |
| | | <18.5 or ≥30 | 0 |
| | Waist circumference (cm)[a] | Men: <94; Women: <80 | 0.5 |
| | | Men: 94 to <102; Women: 80 to <88 | 0.25 |
| | | Men: ≥102; Women: ≥88 | 0 |
| Be physically active | Physical activity (MET-mins/week)[b] | ≥3000 | 1 |
| | | 600 to <3000 | 0.5 |
| | | <600 | 0 |
| Eat a diet rich in wholegrains, vegetables, fruit, and beans | Fruit and vegetable intake (serves/day) | ≥5 | 0.5 |
| | | 3 to <5 | 0.25 |
| | | <3 | 0 |
| | | (1 serve = 1 piece fresh fruit, 5 pieces dried fruit, or 3 heaped tablespoons of cooked or raw vegetables) | |
| | Wholegrain intake (serves/day) | ≥5.5 | 0.5 |
| | | >2 to <5.5 | 0.25 |
| | | ≤2 | 0 |
| | | (1 serve = 1 bowl cereal or 1 slice wholemeal or wholegrain bread. Cereal type not considered due to large amount of missingness. Wholegrain intake calculated without cereal type was slightly more correlated with dietary fibre intake from 24 hr diet recall than wholegrain intake calculated with cereal type considered) | |
| Limit consumption of red and processed meat | Red meat intake (times/week) | <2 times/week | 0.5 |
| | | 2-4 times/week | 0.25 |
| | | >4 times/week | 0 |
| | Processed meat intake (times/week) | <once/week | 0.5 |
| | | once/week | 0.25 |
| | | ≥twice/week | 0 |
| Limit alcohol consumption | Alcohol intake (g/day)[c] | 0.0 | 1 |
| | | Males: >0 to ≤28; Females: >0 to ≤14 | 0.5 |
| | | Males: >28; Females: >14 | 0 |
| | | (1 glass champagne/red wine/white wine = 12.8 g, 1 pint of beer = 18.4 g, 1 measure of spirits (25 ml) = 8 g, 1 standard measure of fortified wine (50 ml) = 8 g; 1 glass of other alcoholic drinks (e.g. alcopops) = 12 g.[1] | |
| Don't smoke | Current Smoking Status | Never Smoker | 1 |
| | | Former Smoker | 0.5 |
| | | Current Smoker | 0 |

[a] Those with a waist circumference (WC) < 50 cm were excluded (i.e. WC recoded to missing; n=7)

[b] Physical activity calculated as metabolic equivalent (MET)-mins/week using IPAQ short form guidelines[2] and the self-reported estimated time spent (on a typical day) doing at least 2 of the 3 following activities – walking, moderate physical activity, and vigorous physical activity.

[c] Estimated from self-reported frequency of alcohol intake and weekly or monthly quantities of different types of alcoholic beverages consumed. For those missing information on quantity of "other alcoholic drinks", intake was estimated from the quantities reported for wine, beer, spirits, and fortified wine (n=224 609). For those who reported consuming alcohol less than weekly and had missing data on quantities of different beverage types, scores were estimated from data available (n=1166) or, where there were no data on quantities consumed, a score of 0.5 was allocated (n=73 137).

#### Table S3 – Definition of additional covariates included in further statistical models

| Cancer Type | Additional covariates included in Model 2 |
|---|---|
| Colorectal cancer | Height (cm), history of inflammatory bowel disease, Crohn's disease, or ulcerative colitis (yes/no) |
| Breast cancer | Height (cm), ever hormone replacement therapy use (yes/no), ever oral contraceptive pill use (yes/no), number of live births, age at onset of menopause (last menstrual period; yes/no), age at menarche (years) |
| Uterine cancer | Height (cm), ever hormone replacement therapy use (yes/no), number of live births, have had menopause at baseline (yes/no), age at menarche (years) |
| Kidney cancer | Height (cm) |
| Lung cancer | History of chronic obstructive pulmonary disease or pulmonary fibrosis (yes/no) |
| Melanoma | Height (cm), time spent outdoors in summer (hours/day), use of sun/UV protection (never/rarely, sometimes, most of the time, always, do not go out in sunshine), solarium use (yes/no) |
| Non-Hodgkin lymphoma | History of any of the following autoimmune diseases - Sjogren syndrome, systemic lupus erythematosus, haemolytic anaemia, rheumatoid arthritis, pernicious anaemia, myasthenia gravis, celiac disease, immune thrombocytopenic purpura, inflammatory bowel disease, Crohn's disease, ulcerative colitis, multiple sclerosis, systemic sclerosis or scleroderma, poly- or dermatomyositis, psoriasis, sarcoidosis, type 1 diabetes (yes/no) |
| Oral cavity/pharyngeal cancer | Time spent outdoors in summer (hours/day), use of sun/UV protection (never/rarely, sometimes, most of the time, always, do not go out in sunshine) |
| Ovarian cancer | Height (cm), ever hormone replacement therapy use (yes/no), ever oral contraceptive pill use (yes/no), number of live births, have had menopausal at baseline (yes/no), age at menarche (years), tubal ligation or fallopian tubes removed (yes/no) |
| Pancreatic cancer | Height (cm) |
| Prostate cancer | Height (cm) |

#### Table S4 - Adjusted associations between the Healthy Lifestyle Index, Polygenic Risk Score, and cancer risk

| | | Healthy lifestyle index | | | | Polygenic risk score | | | |
|---|---|---|---|---|---|---|---|---|---|
| **Cancer Type** | **No. of cases/ person-years** | **High** | **Intermediate** | **Low** | $p_{trend}$ | **Low** | **Intermediate** | **High** | $p_{trend}$ |
| Overall cancer | | | | | | | | | |
| Model 1[a] | 15 240/1 926 987 | Ref | 1.15 (1.11, 1.20) | 1.32 (1.26, 1.37) | <0.0001 | Ref | 1.08 (1.04, 1.13) | 1.20 (1.16, 1.25) | <0.0001 |
| Prostate cancer | | | | | | | | | |
| Model 1[b] | 4476/981 844 | Ref | 0.99 (0.91, 1.06) | 0.92 (0.86, 0.99) | 0.024 | Ref | 1.30 (1.20, 1.40) | 1.89 (1.75, 2.03) | <0.0001 |
| Model 2[c] | 4476/981 844 | Ref | 0.99 (0.91, 1.06) | 0.92 (0.86, 0.99) | 0.024 | Ref | 1.30 (1.20, 1.40) | 1.89 (1.75, 2.03) | <0.0001 |
| Colorectal cancer | | | | | | | | | |
| Model 1[a] | 2150/1 990 006 | Ref | 1.14 (1.02, 1.28) | 1.42 (1.28, 1.59) | <0.0001 | Ref | 1.49 (1.33, 1.67) | 2.04 (1.82, 2.27) | <0.0001 |
| Model 2[d] | 2150/1 990 006 | Ref | 1.14 (1.02, 1.28) | 1.42 (1.27, 1.58) | <0.0001 | Ref | 1.49 (1.33, 1.67) | 2.04 (1.82, 2.27) | <0.0001 |
| Post-menopausal breast cancer | | | | | | | | | |
| Model 1[b] | 1990/576 641 | Ref | 1.36 (1.22, 1.52) | 1.42 (1.27, 1.59) | <0.0001 | Ref | 1.35 (1.20, 1.52) | 1.73 (1.55, 1.94) | <0.0001 |
| Model 2[e] | 1837/537 658 | Ref | 1.34 (1.20, 1.50) | 1.42 (1.26, 1.59) | <0.0001 | Ref | 1.36 (1.20, 1.53) | 1.75 (1.56, 1.96) | <0.0001 |
| Lung cancer | | | | | | | | | |
| Model 1[a] | 1256/1 994 812 | Ref | 1.59 (1.32, 1.93) | 3.50 (2.96, 4.15) | <0.0001 | Ref | 1.30 (1.12, 1.50) | 1.64 (1.43, 1.89) | <0.0001 |
| Model 2[f] | 1256/1 994 812 | Ref | 1.58 (1.30, 1.91) | 3.33 (2.81, 3.94) | <0.0001 | Ref | 1.30 (1.12, 1.50) | 1.64 (1.42, 1.88) | <0.0001 |
| Melanoma | | | | | | | | | |
| Model 1[a] | 1200/1 994 915 | Ref | 1.13 (0.98, 1.30) | 0.98 (0.85, 1.13) | 0.71 | Ref | 1.37 (1.17, 1.60) | 2.03 (1.75, 2.34) | <0.0001 |
| Model 2[g] | 1118/1 844 648 | Ref | 1.14 (0.98, 1.32) | 1.00 (0.86, 1.16) | 0.97 | Ref | 1.39 (1.18, 1.63) | 2.06 (1.77, 2.40) | <0.0001 |
| Non-Hodgkin lymphoma | | | | | | | | | |
| Model 1[a] | 721/1 997 000 | Ref | 0.98 (0.82, 1.18) | 1.01 (0.84, 1.21) | 0.92 | Ref | 1.20 (1.00, 1.45) | 1.42 (1.18, 1.70) | 0.0002 |
| Model 2[h] | 721/1 997 000 | Ref | 0.98 (0.81, 1.18) | 1.00 (0.84, 1.20) | 0.95 | Ref | 1.20 (0.99, 1.45) | 1.42 (1.18, 1.70) | 0.0002 |

| Cancer Type | No. of cases/ person-years | Healthy lifestyle index: High | Intermediate | Low | $p_{trend}$ | Polygenic risk score: Low | Intermediate | High | $p_{trend}$ |
|---|---|---|---|---|---|---|---|---|---|
| Kidney cancer | | | | | | | | | |
| Model 1[a] | 547/1 997 962 | Ref | 1.73 (1.36, 2.21) | 1.91 (1.51, 2.42) | <0.0001 | Ref | 1.18 (0.95, 1.47) | 1.47 (1.20, 1.81) | 0.0002 |
| Model 2[c] | 547/1 997 962 | Ref | 1.73 (1.36, 2.21) | 1.91 (1.51, 2.41) | <0.0001 | Ref | 1.18 (0.95, 1.47) | 1.47 (1.20, 1.81) | 0.0002 |
| Uterine cancer | | | | | | | | | |
| Model 1[b] | 494/995 300 | Ref | 1.31 (1.05, 1.65) | 1.63 (1.31, 2.04) | <0.0001 | Ref | 1.60 (1.26, 2.02) | 1.87 (1.48, 2.36) | <0.0001 |
| Model 2[e] | 466/826 638 | Ref | 1.33 (1.05, 1.67) | 1.63 (1.29, 2.04) | <0.0001 | Ref | 1.65 (1.29, 2.11) | 1.89 (1.48, 2.40) | <0.0001 |
| Pancreatic cancer | | | | | | | | | |
| Model 1[a] | 451/1 998 456 | Ref | 1.54 (1.18, 2.00) | 1.98 (1.54, 2.55) | <0.0001 | Ref | 1.62 (1.25, 2.09) | 2.26 (1.77, 2.89) | <0.0001 |
| Model 2[c] | 451/1 998 456 | Ref | 1.53 (1.18, 2.00) | 1.98 (1.54, 2.54) | <0.0001 | Ref | 1.62 (1.25, 2.10) | 2.26 (1.77, 2.89) | <0.0001 |
| Bladder cancer | | | | | | | | | |
| Model 1[a] | 424/1 998 387 | Ref | 1.23 (0.92, 1.64) | 2.03 (1.57, 2.64) | <0.0001 | Ref | 1.49 (1.16, 1.91) | 1.61 (1.26, 2.06) | 0.0002 |
| Oral cavity/ pharyngeal cancer | | | | | | | | | |
| Model 1[a] | 415/1 998 464 | Ref | 1.24 (0.94, 1.64) | 1.69 (1.31, 2.18) | <0.0001 | Ref | 1.06 (0.84, 1.35) | 1.06 (0.84, 1.35) | 0.60 |
| Model 2[j] | 402/1 949 841 | Ref | 1.21 (0.92, 1.60) | 1.60 (1.23, 2.07) | 0.0002 | Ref | 1.06 (0.83, 1.34) | 1.07 (0.84, 1.36) | 0.59 |
| Ovarian cancer | | | | | | | | | |
| Model 1[b] | 315/996 236 | Ref | 1.13 (0.86, 147) | 1.04 (0.79, 1.38) | 0.75 | Ref | 1.26 (0.95, 1.68) | 1.47 (1.11, 1.94) | 0.0065 |
| Model 2[k] | 282/827 032 | Ref | 1.18 (0.89, 1.56) | 1.03 (0.77, 1.40) | 0.77 | Ref | 1.24 (0.92, 1.68) | 1.45 (1.09, 1.95) | 0.012 |
| Lymphocytic leukaemia | | | | | | | | | |
| Model 1[a] | 246/1 999 253 | Ref | 1.05 (0.76, 1.44) | 0.96 (0.70, 1.32) | 0.78 | Ref | 1.13 (0.78, 1.63) | 2.45 (1.78, 3.36) | <0.0001 |

Overall cancer = Overall incident cases of the 13 cancer types assessed in this study
[a] Cox proportional hazards regression adjusted for age at baseline, sex, assessment centre, 40 principal components of ancestries (PCs), Townsend Index, education, birth location, and income

[b] Cox proportional hazards regression adjusted for age at baseline, assessment centre, 40 PCs, Townsend Index, education, birth location, and income

[c] Cox proportional hazards regression adjusted for covariates in model 1[a] + height

[d] Cox proportional hazards regression adjusted for covariates in model 1[a] + height and history of inflammatory bowel disease
[e] Cox proportional hazards regression adjusted for covariates in model 1[b] + height, ever hormone replacement therapy use, ever oral contraceptive pill use, number of live births, age of menopause, age at menarche

[f] Cox proportional hazards regression adjusted for covariates in model 1[a] + history of chronic obstructive pulmonary disease or pulmonary fibrosis

[g] Cox proportional hazards regression adjusted for covariates in model 1[a] + height, time spent outdoors in summer, use of sun/UV protection, solarium use

[h] Cox proportional hazards regression adjusted for covariates in model 1[a] + history of autoimmune disease
[i] Cox proportional hazards regression adjusted for covariates in model 1[b] + height, ever hormone replacement therapy use, number of live births, menopausal status, age at menarche

[j] Cox proportional hazards regression adjusted for covariates in model 1[a] + time spent outdoors in summer, use of sun/UV protection
[k] Cox proportional hazards regression adjusted for covariates in model 1[b] + height, ever hormone replacement therapy use, ever oral contraceptive pill use, number of live births, menopausal status, age at menarche, tubal ligation or fallopian tubes removed

**Table S5 – Relative excess risk due to interaction (RERI) for additive interactions between Healthy Lifestyle Index and Polygenic Risk Score for overall cancer and 13 cancer types**

| Healthy Lifestyle Index | Polygenic Risk Score | | | | | | Overall test for RERI (p-value) |
|---|---|---|---|---|---|---|---|
| | **Intermediate** | | | **High** | | | |
| | **RERI** | **95%CI** | **p-value** | **RERI** | **95%CI** | **p-value** | |
| **Overall cancer** | | | | | | | 0.049 |
| Intermediate | 0.07 | -0.05, 0.18 | 0.24 | -0.007 | -0.13, 0.11 | 0.91 | |
| Unfavourable | 0.04 | -0.07, 0.16 | 0.45 | 0.08 | -0.04, 0.20 | 0.17 | |
| **Prostate cancer** | | | | | | | 0.14 |
| Intermediate | 0.22 | 0.007, 0.43 | 0.043 | 0.16 | -0.08, 0.41 | 0.20 | |
| Unfavourable | 0.06 | -0.14, 0.26 | 0.57 | -0.09 | -0.33, 0.15 | 0.46 | |
| **Colorectal cancer** | | | | | | | <0.0001 |
| Intermediate | 0.32 | -0.03, 0.67 | 0.071 | 0.46 | 0.08, 0.85 | 0.017 | |
| Unfavourable | 0.25 | -0.11, 0.61 | 0.18 | 0.61 | 0.22, 0.99 | 0.0021 | |
| **Post-menopausal breast cancer** | | | | | | | 0.0001 |
| Intermediate | -0.09 | -0.51, 0.33 | 0.68 | 0.22 | -0.21, 0.65 | 0.32 | |
| Unfavourable | -0.04 | -0.47, 0.39 | 0.86 | 0.43 | -0.02, 0.88 | 0.059 | |
| **Lung cancer** | | | | | | | <0.0001 |
| Intermediate | 0.43 | -0.08, 0.94 | 0.098 | 0.53 | -0.009, 1.08 | 0.054 | |
| Unfavourable | 1.21 | 0.60, 1.82 | 0.0001 | 1.96 | 1.25, 2.67 | <0.0001 | |
| **Melanoma** | | | | | | | 0.40 |
| Intermediate | 0.46 | 0.07, 0.84 | 0.021 | 0.08 | -0.39, 0.55 | 0.74 | |
| Unfavourable | 0.43 | 0.08, 0.78 | 0.016 | 0.25 | -0.19, 0.68 | 0.26 | |
| **Non-Hodgkin lymphoma** | | | | | | | 0.67 |
| Intermediate | 0.59 | 0.21, 0.97 | 0.0023 | 0.20 | -0.26, 0.66 | 0.40 | |

| | | | | | | | |
|---|---|---|---|---|---|---|---|
| Unfavourable | 0.51 | 0.13, 0.89 | 0.0090 | -0.08 | -0.56, 0.39 | 0.74 | |
| **Kidney cancer** | | | | | | | 0.0049 |
| Intermediate | -0.37 | -1.47, 0.72 | 0.50 | -0.34 | -1.46, 0.78 | 0.55 | |
| Unfavourable | -0.12 | -1.11, 0.87 | 0.81 | 0.76 | -0.22, 1.73 | 0.13 | |
| **Uterine cancer** | | | | | | | 0.098 |
| Intermediate | 0.69 | -0.06, 1.44 | 0.073 | 0.14 | -0.74, 1.03 | 0.75 | |
| Unfavourable | 0.92 | 0.06, 1.79 | 0.037 | -0.04 | -1.02, 0.95 | 0.94 | |
| **Pancreatic cancer** | | | | | | | 0.0002 |
| Intermediate | 0.38 | -0.68, 1.44 | 0.48 | 0.48 | -0.68, 1.65 | 0.42 | |
| Unfavourable | 0.70 | -0.34, 1.74 | 0.19 | 1.55 | 0.33, 2.77 | 0.013 | |
| **Bladder cancer** | | | | | | | 0.0002 |
| Intermediate | 0.07 | -0.81, 0.96 | 0.87 | 0.31 | -0.55, 1.17 | 0.48 | |
| Unfavourable | 0.73 | -0.14, 1.59 | 0.099 | 1.12 | 0.25, 1.99 | 0.012 | |
| **Oral cavity/pharyngeal cancer** | | | | | | | 0.73 |
| Intermediate | 0.57 | -0.03, 1.17 | 0.061 | 0.13 | -0.53, 0.79 | 0.69 | |
| Unfavourable | 0.44 | -0.18, 1.06 | 0.17 | 0.10 | -0.58, 0.78 | 0.77 | |
| **Ovarian cancer** | | | | | | | 0.27 |
| Intermediate | 0.08 | -0.62, 0.78 | 0.81 | 0.65 | -0.03, 1.33 | 0.060 | |
| Unfavourable | 0.15 | -0.56, 0.85 | 0.69 | 0.44 | -0.24, 1.12 | 0.21 | |
| **Lymphocytic leukaemia** | | | | | | | 0.69 |
| Intermediate | -0.36 | -1.42, 0.69 | 0.50 | 0.95 | -0.14, 2.04 | 0.086 | |
| Unfavourable | -0.40 | -1.43, 0.64 | 0.45 | 0.49 | -0.53, 1.51 | 0.35 | |

Overall cancer = overall incident cases of the 13 cancer types assessed in this study

Cox proportional hazards regression models adjusted for age at baseline, sex (where relevant), assessment centre, 40 principal components of ancestries, Townsend Index, education, birth location, and income

**References**

1. Alcohol Change UK. Alcohol units. Accessed February 23, 2021. https://alcoholchange.org.uk/alcohol-facts/interactive-tools/check-your-drinking/alcohol-units.

2. The IPAQ Group. Guidelines for the data processing and analysis of the International Physical Activity Questionnaire (IPAQ) – short and long forms. Accessed February 25, 2021. https://www.researchgate.net/file.PostFileLoader.html?id=5641f4c36143250eac8b45b7&assetKey=AS%3A294237418606593%401447163075131.
